# Preliminary Evidence on Long COVID in children

**DOI:** 10.1101/2021.01.23.21250375

**Authors:** Danilo Buonsenso, Daniel Munblit, Cristina De Rose, Dario Sinatti, Antonia Ricchiuto, Angelo Carfi, Piero Valentini

## Abstract

There is increasing evidence that adult patients diagnosed with acute COVID-19 suffer from Long COVID initially described in Italy.

To date, data on Long COVID in children are lacking.

We assessed persistent symptoms in pediatric patients previously diagnosed with COVID-19. More than a half reported at least one persisting symptom even after 120 days since COVID-19, with 42.6% being impaired by these symptoms during daily activities. Symptoms like fatigue, muscle and joint pain, headache, insomnia, respiratory problems and palpitations were particularly frequent, as also described in adults.

The evidence that COVID-19 can have long-term impact children as well, including those with asymptomatic/paucisymptomatic COVID-19, highlight the need for pediatricians, mental health experts and policy makers of implementing measures to reduce impact of the pandemic on child’s health.

## Background

There is increasing evidence that adult patients diagnosed with acute COVID-19 suffer from Long COVID initially described in Italy (1). A recent large cohort of 1733 patients from Wuhan found persistent symptoms in 76% of patients 6 months after initial diagnosis (2). To date, data on Long COVID in children are lacking. We assessed persistent symptoms in pediatric patients previously diagnosed with COVID-19.

## Methods

This cross-sectional study included all children ≤18 year-old diagnosed with microbiologically-confirmed COVID-19 in Fondazione Policlinico Univeersitario A. Gemelli IRCCS (Rome, Italy). Patients > 18 years-old or with severe disability were excluded. Caregivers were interviewed about their child’s health using a questionnaire (supplementary material) developed by the Long Covid ISARIC study group (3), for evaluation of persisting symptoms. Participants were interviewed by two pediatricians, either online or in the outpatient department, from September 1^st^ to January 1^st^. Participants were categorized into groups according to symptoms status during the acute phase (symptomatic/asymptomatic), need for hospitalization and time from COVID-19 diagnosis to follow-up evaluation (<60, 60-120, > 120 days). Numerical variables were compared using t-test or ANOVA, and categorical variables with χ^2^ or Fisher’s exact test where appropriate. All analyses were performed using R version 4.0.3 (R Foundation). This study was approved by the Institutional Ethics Committee and all participants consented to participate.

## Results

129 children diagnosed with COVID-19 between March and November, 2020 were enrolled (mean age of 11 ± 4.4 years, 62 (48.1%) female). Subsequently, three developed Multisystem Inflammatory Syndrome (2.3%) and two myocarditis (1.6%). Patients were assessed on average 162.5 ± 113.7 days after COVID-19 microbiological diagnosis. 41.8% completely recovered, 35.7% had 1 or 2 symptoms and 22.5% had 3 or more. 52.7% had at least one symptom 120 days or more after diagnosis (Table 1).

**Table 1.** Demographic and Clinical Characteristics of the Study Sample (N = 129). PICU: pediatric intensive care unite; MIS-C; multisystem inflammatory syndrome in children.

| **Characteristics** | **Value** |
| --- | --- |
| **age** | 11.0 (4.4) |
| **sex: F** | 62 (48.1%) |
| **ethnicity** |  |
| *white* | 115 (89.1%) |
| *black* | 4 (3.1%) |
| *Latino/hispanic* | 9 (7%) |
| *Arab* | 1 (0.8%) |
| **Comorbidities** |  |
| *Neurological* | 13 (10.1%) |
| *Heart_diseases* | 1 (0.8%) |
| *Asthma* | 5 (3.9%) |
| *Allergic_rhinitis* | 4 (3.1%) |
| *Skin_problems* | 6 (4.7%) |
| *Gut_problems* | 1 (0.8%) |
| *Haematology* | 0 (0%) |
| *Oncology* | 1 (0.8%) |
| *Immune_system_diseases* | 1 (0.8%) |
| *Genetic_conditions* | 1 (0.8%) |
| *Diabetes* | 0 (0%) |
| *Renal_Kidney_problems* | 0 (0%) |
| *Excessive_weight_obesity* | 3 (2.3%) |
| *Rheumatology* | 1 (0.8%) |
| **Symtomatic** | 96 (74.4%) |
| **Hospitalized** | 6 (4.7%) |
| **Admitted to PICU** | 3 (2.3%) |
| **Diagnosed with any of the following conditions AFTER acute COVID-19** |  |
| *MIS-C* | 3 (2.3%) |
| *Pulmonary embolism* | 0 (0%) |
| *Myocarditis* | 2 (1.6%) |
| *Asthma* | 3 (2.3%) |
| *Coagulopathy* | 0 (0%) |
| *Kidney problems* | 0 (0%) |
| *Type 1 Diabetes* | 0 (0%) |
| **Distance from diagnosis of acute COVID-19 (days)** | 162.5 (113.7) |
| **Persistent symptoms** |  |
| *None* | 54 (41.9%) |
| *1 or 2* | 46 (35.7%) |
| *3 or more* | 29 (22.5%) |
| **Any persisting symptom at follow-up** |  |
| *< 60 days* | 31 (24%) |
| *60-119 days* | 30 (23.3%) |
| *120 or more days* | 68 (52.7%) |
| **QoL Before COVID-19** | 96.3 (5.3) |
| **QoL at time of survey** | 92.9 (9.1) |

Table 2 provides details about persistence of symptoms according to severity and length of follow-up. Insomnia (18.6%), respiratory symptoms (including pain and chest tightness) (14.7%), nasal congestion (12.4%), fatigue (10.8%), muscle (10.1%) and joint pain (6.9%), and concentration difficulties (10.1), were the most frequently reported symptoms. Although they were more common in symptomatic or hospitalized children, they were also described in those individuals who were asymptomatic during acute phase. 29 out of the 68 (42.6%) children assessed ≥120 days from diagnosis were still distressed by these symptoms.

**Table 2.**
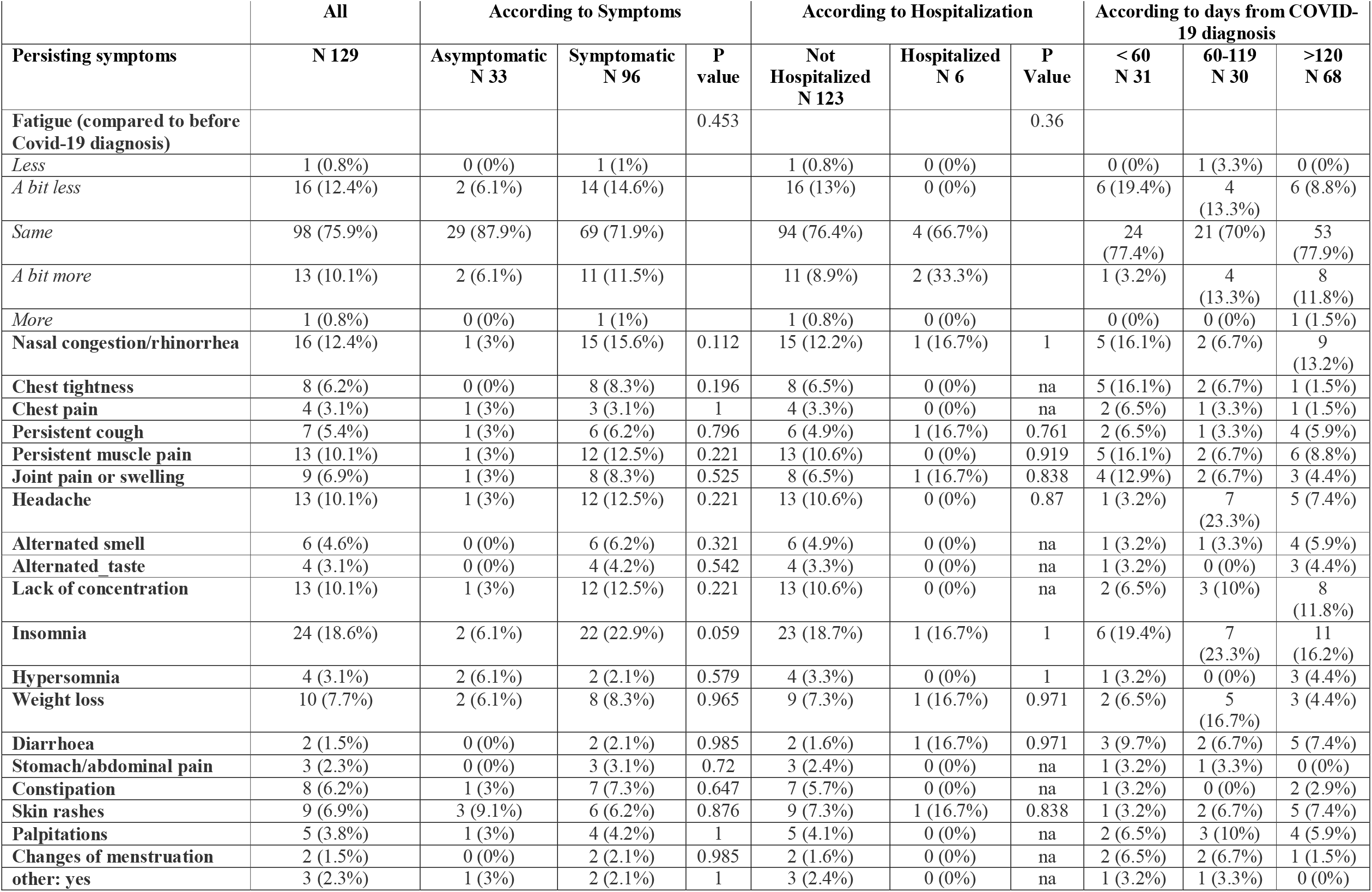
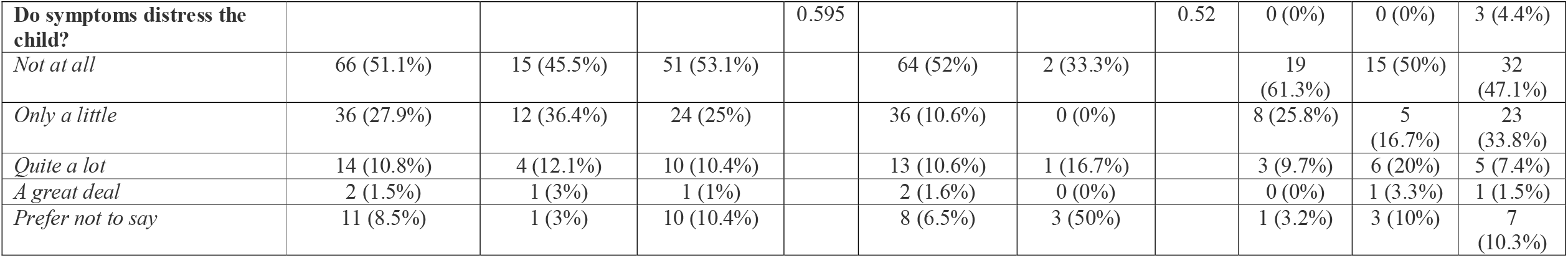
Persisting symptoms in children with COVID-19, according to symptoms, need of hospitalization and distance from diagnosis of acute COVID-19. NA: not applicable. Others include: hair loss (n 2) and skin peeling (n 1)

|  | All | According to Symptoms |  |  | According to Hospitalization |  |  | According to days from COVID-19 diagnosis |  |  |
| --- | --- | --- | --- | --- | --- | --- | --- | --- | --- | --- |
| Persisting symptoms | N 129 | Asymptomatic<br>N 33 | Symptomatic<br>N 96 | P<br>value | Not<br>Hospitalized<br>N 123 | Hospitalized<br>N 6 | P<br>Value | < 60<br>N 31 | 60-119<br>N 30 | >120<br>N 68 |
| Fatigue (compared to before Covid-19 diagnosis) |  |  |  | 0.453 |  |  | 0.36 |  |  |  |
| *Less* | 1 (0.8%) | 0 (0%) | 1 (1%) |  | 1 (0.8%) | 0 (0%) |  | 0 (0%) | 1 (3.3%) | 0 (0%) |
| *A bit less* | 16 (12.4%) | 2 (6.1%) | 14 (14.6%) |  | 16 (13%) | 0 (0%) |  | 6 (19.4%) | 4<br>(13.3%) | 6 (8.8%) |
| *Same* | 98 (75.9%) | 29 (87.9%) | 69 (71.9%) |  | 94 (76.4%) | 4 (66.7%) |  | 24<br>(77.4%) | 21 (70%) | 53<br>(77.9%) |
| *A bit more* | 13 (10.1%) | 2 (6.1%) | 11 (11.5%) |  | 11 (8.9%) | 2 (33.3%) |  | 1 (3.2%) | 4<br>(13.3%) | 8<br>(11.8%) |
| *More* | 1 (0.8%) | 0 (0%) | 1 (1%) |  | 1 (0.8%) | 0 (0%) |  | 0 (0%) | 0 (0%) | 1 (1.5%) |
| Nasal congestion/rhinorrhea | 16 (12.4%) | 1 (3%) | 15 (15.6%) | 0.112 | 15 (12.2%) | 1 (16.7%) | 1 | 5 (16.1%) | 2 (6.7%) | 9<br>(13.2%) |
| Chest tightness | 8 (6.2%) | 0 (0%) | 8 (8.3%) | 0.196 | 8 (6.5%) | 0 (0%) | na | 5 (16.1%) | 2 (6.7%) | 1 (1.5%) |
| Chest pain | 4 (3.1%) | 1 (3%) | 3 (3.1%) | 1 | 4 (3.3%) | 0 (0%) | na | 2 (6.5%) | 1 (3.3%) | 1 (1.5%) |
| Persistent cough | 7 (5.4%) | 1 (3%) | 6 (6.2%) | 0.796 | 6 (4.9%) | 1 (16.7%) | 0.761 | 2 (6.5%) | 1 (3.3%) | 4 (5.9%) |
| Persistent muscle pain | 13 (10.1%) | 1 (3%) | 12 (12.5%) | 0.221 | 13 (10.6%) | 0 (0%) | 0.919 | 5 (16.1%) | 2 (6.7%) | 6 (8.8%) |
| Joint pain or swelling | 9 (6.9%) | 1 (3%) | 8 (8.3%) | 0.525 | 8 (6.5%) | 1 (16.7%) | 0.838 | 4 (12.9%) | 2 (6.7%) | 3 (4.4%) |
| Headache | 13 (10.1%) | 1 (3%) | 12 (12.5%) | 0.221 | 13 (10.6%) | 0 (0%) | 0.87 | 1 (3.2%) | 7<br>(23.3%) | 5 (7.4%) |
| Alternated smell | 6 (4.6%) | 0 (0%) | 6 (6.2%) | 0.321 | 6 (4.9%) | 0 (0%) | na | 1 (3.2%) | 1 (3.3%) | 4 (5.9%) |
| Alternated_taste | 4 (3.1%) | 0 (0%) | 4 (4.2%) | 0.542 | 4 (3.3%) | 0 (0%) | na | 1 (3.2%) | 0 (0%) | 3 (4.4%) |
| Lack of concentration | 13 (10.1%) | 1 (3%) | 12 (12.5%) | 0.221 | 13 (10.6%) | 0 (0%) | na | 2 (6.5%) | 3 (10%) | 8<br>(11.8%) |
| Insomnia | 24 (18.6%) | 2 (6.1%) | 22 (22.9%) | 0.059 | 23 (18.7%) | 1 (16.7%) | 1 | 6 (19.4%) | 7<br>(23.3%) | 11<br>(16.2%) |
| Hypersomnia | 4 (3.1%) | 2 (6.1%) | 2 (2.1%) | 0.579 | 4 (3.3%) | 0 (0%) | 1 | 1 (3.2%) | 0 (0%) | 3 (4.4%) |
| Weight loss | 10 (7.7%) | 2 (6.1%) | 8 (8.3%) | 0.965 | 9 (7.3%) | 1 (16.7%) | 0.971 | 2 (6.5%) | 5<br>(16.7%) | 3 (4.4%) |
| Diarrhoea | 2 (1.5%) | 0 (0%) | 2 (2.1%) | 0.985 | 2 (1.6%) | 1 (16.7%) | 0.971 | 3 (9.7%) | 2 (6.7%) | 5 (7.4%) |
| Stomach/abdominal pain | 3 (2.3%) | 0 (0%) | 3 (3.1%) | 0.72 | 3 (2.4%) | 0 (0%) | na | 1 (3.2%) | 1 (3.3%) | 0 (0%) |
| Constipation | 8 (6.2%) | 1 (3%) | 7 (7.3%) | 0.647 | 7 (5.7%) | 0 (0%) | na | 1 (3.2%) | 0 (0%) | 2 (2.9%) |
| Skin rashes | 9 (6.9%) | 3 (9.1%) | 6 (6.2%) | 0.876 | 9 (7.3%) | 1 (16.7%) | 0.838 | 1 (3.2%) | 2 (6.7%) | 5 (7.4%) |
| Palpitations | 5 (3.8%) | 1 (3%) | 4 (4.2%) | 1 | 5 (4.1%) | 0 (0%) | na | 2 (6.5%) | 3 (10%) | 4 (5.9%) |
| Changes of menstruation | 2 (1.5%) | 0 (0%) | 2 (2.1%) | 0.985 | 2 (1.6%) | 0 (0%) | na | 2 (6.5%) | 2 (6.7%) | 1 (1.5%) |
| other: yes | 3 (2.3%) | 1 (3%) | 2 (2.1%) | 1 | 3 (2.4%) | 0 (0%) | na | 1 (3.2%) | 1 (3.3%) | 0 (0%) |
| **Do symptoms distress the child?** |  |  |  | 0.595 |  |  | 0.52 | 0 (0%) | 0 (0%) | 3 (4.4%) |
| *Not at all* | 66 (51.1%) | 15 (45.5%) | 51 (53.1%) |  | 64 (52%) | 2 (33.3%) |  | 19 (61.3%) | 15 (50%) | 32 (47.1%) |
| *Only a little* | 36 (27.9%) | 12 (36.4%) | 24 (25%) |  | 36 (10.6%) | 0 (0%) |  | 8 (25.8%) | 5 (16.7%) | 23 (33.8%) |
| *Quite a lot* | 14 (10.8%) | 4 (12.1%) | 10 (10.4%) |  | 13 (10.6%) | 1 (16.7%) |  | 3 (9.7%) | 6 (20%) | 5 (7.4%) |
| *A great deal* | 2 (1.5%) | 1 (3%) | 1 (1%) |  | 2 (1.6%) | 0 (0%) |  | 0 (0%) | 1 (3.3%) | 1 (1.5%) |
| *Prefer not to say* | 11 (8.5%) | 1 (3%) | 10 (10.4%) |  | 8 (6.5%) | 3 (50%) |  | 1 (3.2%) | 3 (10%) | 7 (10.3%) |

## Discussion

To our knowledge, this is the first study providing evidence of Long Covid in children. More than a half reported at least one persisting symptom even after 120 days since COVID-19, with 42.6% being impaired by these symptoms during daily activities. Symptoms like fatigue, muscle and joint pain, headache, insomnia, respiratory problems and palpitations were particularly frequent, as also described in adults (1, 2).

Limitations of the study include the single-center design with a relatively small sample size. All patients were interviewed once, however this study is designed to continue periodic assessments until 24 months after COVID-19 diagnosis and to include household members of different age groups, with or without COVID-19 as a control group.

Children have been mostly overlooked during this pandemic, since the clinical course of COVID-19 in this group is much milder than in adults (4). However, there is an increasing evidence that restrictive measures aimed at limiting the pandemic are having a significant impact on child’s mental health (5). Childhood is a delicate and fundamental period of life, critical for acquisition of social, behavioral and educational development. The evidence that COVID-19 can have long-term impact children as well, including those with asymptomatic/paucisymptomatic COVID-19, highlight the need for pediatricians, mental health experts and policy makers of implementing measures to reduce impact of the pandemic on child’s health.

## Data Availability

all data are available

