## supplementary material for "Preliminary Evidence on Long COVID in children"

COVID-19 Health and Wellbeing **Initial** Follow Up Survey for Children

**PARENT REPORT FOR CHILDREN AND YOUNG PEOPLE**

**(between 0 and 18 years of age)**

**The question on our minds**

This is for people like you, whose child or children have had Covid-19. We would like your help to answer a question that is on our minds and may be on yours: “What does Covid-19 mean for the long-term health and well-being of your child?”

**How you can help**

This is a new illness. Being included in this survey means you can help us build a better understanding of the acute and long term care and support needed for Covid-19 for children globally. As far as possible, we do not want to leave anyone out. Our aim is that every child with Covid-19 has a chance to take part, whether they have been treated in hospital or at home. We do not know how long symptoms in children will last, so, to find out, we would like to repeat this survey asking you about your child’s health in three to six months’ time after the first symptoms appearance.

**Protecting your information**

In this follow-up survey, we will use information that you provide. However, we will only use information that we need to inform our research. We will let very few people know your and your child’s name or contact details, and only then if they really need it for this study. Everyone involved in this study will keep your data safe and secure. We will also follow all privacy rules.

We will make sure no one can work out who you or your child are from any reports. Your answers will only be used in ways that do not identify you or your child, for example in summary reports or scientific journals to educate health care professionals. Please feel free to read more about the privacy policy at our website, where you can also download the online version or further paper copies of this form.

**Our thanks to you, and an offer**

Thank you for helping answer this important question. Once you have completed the survey, we would also like to offer you the chance to tell us more via a consultation with a nurse, doctor or researcher. We will not be able to do this for everyone, but if you would like the chance to be included, please fill in your and your child’s contact details at the end of the survey.

SURVEY TIMEPOINT (to be completed by the team before sending or administering the survey): 3m [	] 6m [	] 12m [	] 24m [	] 36m [	]

Survey completed: □ Self-assessment □ Staff/research led assessment

□ Online □Telephone □Post □Clinic

□ Mother/female caregiver □ Father/male caregiver □ Other If *other*, please specify ________________

**Your permission to proceed**

Thank you for coming this far. Now to take part, please read the statements below, and initial the boxes if you are happy to go ahead.

| PLEASE MARK YOUR INITIALS AGAINST EACH STATEMENT WITH WHICH YOU AGREE: | *Add your Initials or tick the box:* | |
| --- | --- | --- |
| I give my consent for the information I provide in this study to be used as advised. |  |  |
| I would like to continue to be sent this survey via email, post or to be contacted via telephone follow up every 3 to 6 months for a maximum of 3 years after my child’s Covid-19 illness.  If yes, please enter your contact details here:  E-mail: ___________________________________  Mobile phone number: _______________________  Home telephone number: _____________________ | YES | NO |
| I would like the possibility to be contacted by a nurse, doctor or researcher to discuss my child’s Covid-19 illness further.  If yes, please enter your contact details here:  Mobile phone number: ______________________  Home telephone number: ____________________ | YES | NO |
| You are completing this survey on behalf of your child, please enter your and their details  **Your child’s first name: Surname: _______________**  **Your first name:** **Surname:**  **Town/City of residence: Postcode: ________________**  **Your signature:** | | |

**Local hospital ID:**

**1a.** **About you child**

**Sex assigned at Birth:** □ Male □ Female □ Non-binary □ Prefer not to say

*Ethnicity (tick all that apply):* □ White □ Arab □ Black □ East Asian □ South Asian

□ West Asian □ Latin American □Other (specify):____________□ Prefer not to say

**What is your child’s estimated height:** (□ Cm □ metres □ feet/inches) □ Not sure

**What is your child’s current estimated weight:** (□ kg □ Ibs □ stone) □ Not sure

**What was your child's estimated weight before Covid19 illness?** (□ kg □ Ibs□ stone) □ Not sure

**How many other members regularly live in your household, including yourself:** [_Number_]

**How many years formal school education has your child had?* [_Number_]**

****including primary school (e.g. from around 6 years depending on country)***

**1b.** **About your child’s Covid-19 illness - all the questions relate to his/her health and wellbeing)**

**Date you completed the survey (DD/MM/YYYY):**[_D_][_D_]/[_M_][_M_]/[_2_][_0_][_Y_][_Y_]

**What is your child’s date of birth (DD/MM/YYYY):**[_D_][_D_]/[_M_][_M_]/[_Y_][_Y_][_Y_][_Y_]

**Approximately, what day did you first notice your child was experiencing symptoms of**

**Covid-19?** [_D_][_D_]/[_M_][_M_]/[_2_][_0_][_Y_][_Y_]

**How was your child diagnosed with Covid-19?**

□ Laboratory confirmed (PCR or/and Antibody test)

□ Physician confirmed (no laboratory testing was performed) □ Not sure

**Has your child been admitted to hospital due to Covid-19?**	□ Yes	□ No

*(If the answer is “no”, please, move on to the section “2”; if the answer is “yes”, please, proceed with the following questions)*

- **Roughly at what date was your child first admitted to hospital?** [_D_][_D_]/[_M_][_M_]/[_2_][_0_][_Y_][_Y_]
- **Roughly at what date was your child first discharged from hospital?** [_D_][_D_]/[_M_][_M_]/[_2_][_0_][_Y_][_Y_]
- **Spent any time in the Paediatric Intensive Care Unit (PICU)?** □ Yes □ No □ Not sure
- **Hospital admission after the first acute Covid-19 illness?** □ Yes	□ No

If yes, how many times: [_Number_]

If yes, specify reason/reasons:

______________________________________________________________________________

Name of hospital/hospitals:

- **Visit to other health facility after the first acute Covid-19 illness? □ Yes □ No**

If yes, how many times: [_Number_]

If yes, specify reason/reasons:

______________________________________________________________________________

**2a. About your child’s emotional wellbeing, social relationships and activities’**

To answer the following questions, please **mark an X** on the lines below that shows your opinion on the question. In each scale, **1 is the least favorable value, 5 is the most favorable value.**

**Compared to before your child’s Covid-19 infection, how much is the child now doing the following**

***If there are changes, please indicate whether you think these are due to the illness itself or to the Covid-19 pandemic (e.g. changes in social activities)***

***Eating***

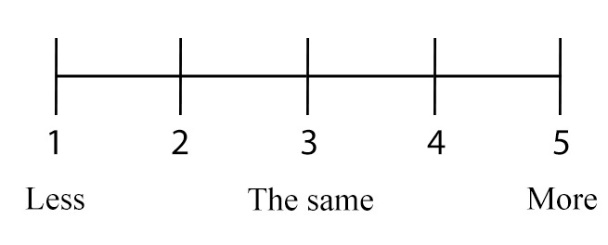

***Fatigue***

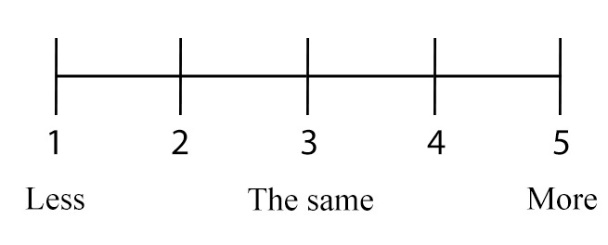

***If there are changes,*** *please indicate*

*whether you think these are due to*

*□ Illness itself □ Covid-19 pandemic □ Both □ Unsure*

• **Do the the current sympyoms upset or distress your child?**

□ Not at all □ Only a little □ Quite a lot □ A great deal

**2b. About your child’s current state of health**

**Has your child been physician’s diagnosed or received treatment for any of the following chronic medical conditions prior to the Covid-19 infection? (answer with a tick in the box)**

|  | **Yes** | **No** | **Unknown** |
| --- | --- | --- | --- |
| Prematurity *(baby born <37 weeks)* |  |  |  |
| Neurological *(pertaining to the nervous system)* |  |  |  |
| Neurodisability |  |  |  |
| Heart diseases |  |  |  |
| Respiratory diseases (not including asthma) |  |  |  |
| Asthma (doctor’s diagnosed) |  |  |  |
| Allergic rhinitis/hay fever |  |  |  |
| Food allergy |  |  |  |
| Atopic dermatitis/Eczema |  |  |  |
| Other skin problems (not including eczema) |  |  |  |
| Gut problems |  |  |  |
| Haematology *(blood diseases)* |  |  |  |
| Oncology *(cancer or other progressively enlarging and spreading*  *tumor)* |  |  |  |
| Immune system diseases | □ |  |  |
| Genetic conditions |  |  |  |
| Diabetes indicate type: □ Type 1 □ Type 2 |  |  |  |
| Other endocrine illness (not diabetes) |  |  |  |
| Renal/Kidney problems |  |  |  |
| Excessive weight and obesity |  |  |  |
| Malnutrition *(deficiencies, excesses, or imbalances in a person's*  *intake of energy and/or nutrients)* |  |  |  |
| Rheumatology *(e.g. arthritis, or inflammation of the joints)* |  |  |  |
| HIV |  |  |  |
| Other (please indicate) |  | | |

**Prior to COVID-19 infection, how was your child’s health in general?**

□ Very poor □ Poor □ Fair (ok) □ Good □ Very good

***If you ticked bad or very bad, please explain:***

**Prior to COVID-19 infection, how would you describe your child’s mental health in general**

□ Very bad □ Bad □ Fair (ok) □ Good □ Very good

***If you ticked bad or very bad, please explain:***

____________________________________________________________________________________

**Has your child ever been under Child and Adolescent Mental Health services before the Covid-19 pandemic?** □ Yes □ No □ Not sure

**Have you requested help because of Covid-19 consequences to your child’s physical health?** □ □ Yes □ No □ Not sure

**Has your child felt feverish recently?** □ Yes □ No □ Not sure

*(If the answer is “no”, please, move on to the next question)*
□ Within the last 7 days □ 1-2 weeks □ >2-4 weeks □ >1-2 months □ >2-3 months □ >3-6 months

□ Since discharge

***If yes, what was the most likely cause of your child’s most recent feverish illness?***

□ Covid-19 □ Other respiratory infection (cough/cold/sore throat) □ TB

□ Stomach infection (diarrhea/vomiting)	□ Urinary infection

□ Other: specify: ___________________________________

□ Unknown □ Prefer not to say

**Date of last positive SARS-CoV-2 /Covid-19 test** [_D_][_D_]/[_M_][_M_]/[_2_][_0_][_Y_][_Y_]

**How much do you agree with the following statement?**
“My child has fully recovered from Covid-19”

Please **mark an X** on the line below that shows your opinion on the question as of **TODAY**

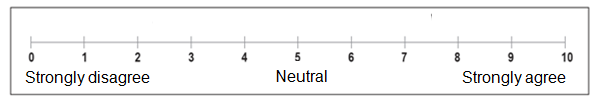

**3. Since having Covid-19, has your child been diagnosed with any of the following?**

| Multisystem inflammatory syndrome | □ Yes | □ No | Shock / Toxic shock syndrome | □ Yes | □ No |
| --- | --- | --- | --- | --- | --- |
| Pulmonary embolism  *(PE, “Clot in lung”)* | □ Yes | □ No | Coagulopathy *(excessive bleeding or clotting)* | □ Yes | □ No |
| Kawasaki disease | □ Yes | □ No | Kidney problems | □ Yes | □ No |
| Respiratory failure | □ Yes | □ No | Type 1 Diabetes | □ Yes | □ No |
| Myocarditis  *(inflammation of the heart muscle)* | □ Yes | □ No | Type 2 Diabetes | □ Yes | □ No |
| Asthma | □ Yes | □ No | Intussusception | □ Yes | □ No |

Other condition (please specify)?

| **4. Within the last seven days, have you had any of these symptoms, which were NOT present prior to Covid-19?**  **If yes, please indicate duration of the symptom(-s).** |
| --- |

| **Respiratory problems** | **Yes/No** | **If yes**, what is the duration of symptoms |  |  |
| --- | --- | --- | --- | --- |
| Nasal congestion / rhinorrhea | □ Yes □ No | □ less than 1 month □ 1-3 months □ over 3 to 6 months □ more than 6 months |  |  |
| Difficulty breathing /chest tightness | □ Yes □ No | □ less than 1 month □ 1-3 months □ over 3 to 6 months □ more than 6 months |  |  |
| Pain on breathing | □ Yes □ No | □ less than 1 month □ 1-3 months □ over 3 to 6 months □ more than 6 months |  |  |
| Chest pain | □ Yes □ No | □ less than 1 month □ 1-3 months □ over 3 to 6 months □ more than 6 months |  |  |
| Persistent cough | □ Yes □ No | □ less than 1 month □ 1-3 months □ over 3 to 6 months □ more than 6 months |  |  |
| *If yes, □ dry cough □ with phlegm* | | |  |  |
| **Musculoskeletal problems** | **Yes/No** | **If yes**, what is the duration of symptoms |  |  |
| Cannot fully move or control movement | □ Yes □ No | □ less than 1 month □ 1-3 months □ over 3 to 6 months □ more than 6 months |  |  |
| Problems with balance | □ Yes □ No | □ less than 1 month □ 1-3 months □ over 3 to 6 months □ more than 6 months |  |  |
| Persistent muscle pain | □ Yes □ No | □ less than 1 month □ 1-3 months □ over 3 to 6 months □ more than 6 months |  |  |
| Joint pain or swelling | □ Yes □ No | □ less than 1 month □ 1-3 months □ over 3 to 6 months □ more than 6 months |  |  |
| **Neurological problems** | **Yes/No** | **If yes**, what is the duration of symptoms |  |  |
| Headache | □ Yes □ No | □ less than 1 month □ 1-3 months □ over 3 to 6 months □ more than 6 months |  |  |
| Dizziness/ light headedness | □ Yes □ No | □ less than 1 month □ 1-3 months □ over 3 to 6 months □ more than 6 months |  |  |
| Fainting/ blackouts | □ Yes □ No | □ less than 1 month □ 1-3 months □ over 3 to 6 months □ more than 6 months |  |  |
| Problems seeing/blurred vision | □ Yes □ No | □ less than 1 month □ 1-3 months □ over 3 to 6 months □ more than 6 months |  |  |
| Disturbed smell | □ Yes □ No | □ less than 1 month □ 1-3 months □ over 3 to 6 months □ more than 6 months |  |  |
| Loss of smell | □ Yes □ No | □ less than 1 month □ 1-3 months □ over 3 to 6 months □ more than 6 months |  |  |
| Disturbed taste | □ Yes □ No | □ less than 1 month □ 1-3 months □ over 3 to 6 months □ more than 6 months |  |  |
| Loss of taste | □ Yes □ No | □ less than 1 month □ 1-3 months □ over 3 to 6 months □ more than 6 months |  |  |
| Tremor/shakiness | □ Yes □ No | □ less than 1 month □ 1-3 months □ over 3 to 6 months □ more than 6 months |  |  |
| Tingling feeling/ “pins and needles“ | □ Yes □ No | □ less than 1 month □ 1-3 months □ over 3 to 6 months □ more than 6 months |  |  |
| Seizures/fits | □ Yes □ No | □ less than 1 month □ 1-3 months □ over 3 to 6 months □ more than 6 months |  |  |
| Confusion/lack of concentration | □ Yes □ No | □ less than 1 month □ 1-3 months □ over 3 to 6 months □ more than 6 months |  |  |
| Problems speaking or communicating | □ Yes □ No | □ less than 1 month □ 1-3 months □ over 3 to 6 months □ more than 6 months |  |  |
| Insomnia *(hard to fall asleep, hard to stay asleep)* | □ Yes □ No | □ less than 1 month □ 1-3 months □ over 3 to 6 months □ more than 6 months |  |  |
| Hypersomnia *(excessive daytime sleepiness or prolonged nighttime sleep)* | □ Yes □ No | □ less than 1 month □ 1-3 months □ over 3 to 6 months □ more than 6 months |  |  |
| **Fatigue** | □ Yes □ No | □ less than 1 month □ 1-3 months □ over 3 to 6 months □ more than 6 months |  |  |
| **Gastrointestinal problems** | **Yes/No** | **If yes**, what is the duration of symptoms |  |  |
| Weight loss | □ Yes □ No | □ less than 1 month □ 1-3 months □ over 3 to 6 months □ more than 6 months |  |  |
| Problems swallowing or chewing | □ Yes □ No | □ less than 1 month □ 1-3 months □ over 3 to 6 months □ more than 6 months |  |  |
| Poor appetite | □ Yes □ No | □ less than 1 month □ 1-3 months □ over 3 to 6 months □ more than 6 months |  |  |
| Diarrhea | □ Yes □ No | □ less than 1 month □ 1-3 months □ over 3 to 6 months □ more than 6 months |  |  |
| Stomach/ abdominal pain | □ Yes □ No | □ less than 1 month □ 1-3 months □ over 3 to 6 months □ more than 6 months |  |  |
| Feeling nauseous | □ Yes □ No | □ less than 1 month □ 1-3 months □ over 3 to 6 months □ more than 6 months |  |  |
| Vomiting | □ Yes □ No | □ less than 1 month □ 1-3 months □ over 3 to 6 months □ more than 6 months |  |  |
| Constipation | □ Yes □ No | □ less than 1 month □ 1-3 months □ over 3 to 6 months □ more than 6 months |  |  |
| **Cardiovascular problems** | **Yes/No** | **If yes**, what is the duration of symptoms |  |  |
| Palpitations (heart racing) | □ Yes □ No | □ less than 1 month □ 1-3 months □ over 3 to 6 months □ more than 6 months |  |  |
| Variations in heart rate (tachycardia or bradycardia) | □ Yes □ No | □ less than 1 month □ 1-3 months □ over 3 to 6 months □ more than 6 months |  |  |
| Bleeding | □ Yes □ No | □ less than 1 month □ 1-3 months □ over 3 to 6 months □ more than 6 months |  |  |
| *If yes, specify bleeding site:* |  |  |  |  |
| **Genitourinary problems** | **Yes/No** | **If yes**, what is the duration of symptoms |  | **If yes**, what is the duration of symptoms |
| Urination problems | □ Yes □ No | □ less than 1 month □ 1-3 months □ over 3 to 6 months □ more than 6 months |  |  |
| Changes in menstruation,  if settled before Covid-19 | □ Yes □ No □ Not applicable | □ less than 1 month □ 1-3 months □ over 3 to 6 months □ more than 6 months |  |  |
| **Other problems** | **Yes/No** | **If yes**, what is the duration of symptoms |  |  |
| Bilateral conjunctivitis | □ Yes □ No | □ less than 1 month □ 1-3 months □ over 3 to 6 months □ more than 6 months |  |  |
| *If yes, □ purulent □ non-purulent* |  |  |  |  |
| Lumps or rashes (purple/pink) on toes | □ Yes □ No | □ less than 1 month □ 1-3 months □ over 3 to 6 months □ more than 6 months |  |  |
| Skin rash  *If yes, tick all body areas that apply* | □ Yes □ No | □ less than 1 month □ 1-3 months □ over 3 to 6 months □ more than 6 months |  |  |
| *Face* | □ Yes □ No | □ less than 1 month □ 1-3 months □ over 3 to 6 months □ more than 6 months |  |  |
| *Trunk (stomach or back)* | □ Yes □ No | □ less than 1 month □ 1-3 months □ over 3 to 6 months □ more than 6 months |  |  |
| *Arms* | □ Yes □ No | □ less than 1 month □ 1-3 months □ over 3 to 6 months □ more than 6 months |  |  |
| *Legs* | □ Yes □ No | □ less than 1 month □ 1-3 months □ over 3 to 6 months □ more than 6 months |  |  |
| *Buttocks* | □ Yes □ No | □ less than 1 month □ 1-3 months □ over 3 to 6 months □ more than 6 months |  |  |
| *Toes* | □ Yes □ No | □ less than 1 month □ 1-3 months □ over 3 to 6 months □ more than 6 months |  |  |
| *Fingers* | □ Yes □ No | □ less than 1 month □ 1-3 months □ over 3 to 6 months □ more than 6 months |  |  |
| *Accompanied by itch* | □ Yes □ No | □ less than 1 month □ 1-3 months □ over 3 to 6 months □ more than 6 months |  |  |
| Other New Symptoms | □ Yes □ No | □ less than 1 month □ 1-3 months □ over 3 to 6 months □ more than 6 months |  |  |

If your child had experienced/is experiencing any **other NEW symptoms** that were not covered by the tables above, please, specify them here. Please also indicate how long did these symptoms last (eg days, weeks, months or they are ongoing). **When did these symptoms start:**

___________________________________________________________________________________________________________________

We would like to know how good or bad your child’s health was

**BEFORE COVID-19** and is **TODAY**

This scale is numbered from 0 to 100%

with **100% meaning the best health** you can imagine

**0% means the worst health** you can imagine.

Please **write the number in the box below each scale** to indicate how good or bad your child’s health was **BEFORE COVID-19** and is **TODAY**.

100

95

90

85

80

75

70

65

60

55

50

45

40

35

30

25

20

15

10

5

0

**Before COVID**

100

95

90

85

80

75

70

65

60

55

50

45

40

35

30

25

20

15

10

5

0

**Today**

| **Please let us know of any further comments about the child’s illness, the pandemic, lockdown and/or any sequelae.** |
| --- |
| **End of survey** |
| **Thank you for your time!** |
